# Estimation of Tunisia COVID-19 infected cases based on mortality rate

**DOI:** 10.1101/2020.04.15.20065532

**Authors:** Ines Abdeljaoued-Tej, Marc Dhenain

**Affiliations:** BIMS Laboratory, LR16IPT09, Institut Pasteur de Tunis, University of Tunis El Manar, Tunisia; Ecole Supérieure de la Statistique et de l’Analyse de l’Information, Université de Carthage, Tunisie; Académie Vétérinaire de France, 34, rue Bréguet, 75011 Paris, France; Académie Nationale de Médecine, 16 rue Bonaparte, 75006 Paris, France; Centre National de la Recherche Scientifique (CNRS), Université Paris-Sud, Université Paris-Saclay UMR 9199, Laboratoire des Maladies Neurodégénératives, 18 Route du Panorama, F-92265 Fontenay-aux-Roses, France; Commissariat à l’Energie Atomique et aux Energies Alternatives (CEA), Institut François Jacob, Molecular Imaging Research Center (MIRCen), 18 Route du Panorama, F-92265 Fontenay-aux-Roses, France

**Keywords:** COVID-19, corona virus, estimated number of cases, mortality, reported prevalence, reported and unreported cases, isolation, quarantine, public closings, epidemic mathematical model

## Abstract

Estimating the number of people affected by COVID-19 is crucial in deciding which public health policies to follow. The authorities in different countries carry out mortality counts. We propose that the mortality reported in each country can be used to create an index of the number of actual cases at a given time. The specificity of whether or not deaths are rapid or not by COVID-19 also affects the number of actual cases. The number of days between the declaration of illness and death varies between 12 and 18 days. For a delay of 18 days, and using an estimated mortality rate of 2%, the number of cases in April 2020 in Tunisia would be ^5 580^ people. The pessimistic scenario predicts ^22 320^ infected people, and the most optimistic predicts 744 (which is the number of reported cases on April ^12, 2020^). Modeling the occurrence of COVID-19 cases is critical to assess the impact of policies to prevent the spread of the virus.

## 1 INTRODUCTION

Infection with the Sars CoV-2 coronavirus that causes COVID-19 has spread throughout the world and is causing a significant number of deaths [12]. Every day estimates of the number of people affected and a death toll are provided by authorities in different countries and distributed worldwide. We address the following fundamental issues concerning this epidemic: How will the epidemic evolve in Tunisia for the number of reported cases and unreported cases? How will the number of unreported cases influence the severity of the epidemic? How will public health measures, such as isolation, and public closings, impact the final size of the epidemic?

Knowing the number of people affected is important for implementing strategies to protect populations and to end the crisis: In [7] the median incubation period of COVID-19 was studied, but publicly reported cases may over-represent severe cases. This number reveals flagrant differences between countries. For example, on April ^13*th*^, a day when the number of worldwide deaths was close to 120 000, Tunisia reported around 707 people affected (60 per million the population) and 31 deaths, compared with many more 136 779 affected persons in France (2 095 per million) for 14 967 deaths. China reported around 82 000 people affected (57 per million) and more than 3 300 deaths. Germany, for its part, reported 130 000 people affected (1 552 per million) and 3 194 deaths. At first glance, differences between countries can be objectified by calculating the mortality rate *T* on a given day (*t*_0_):

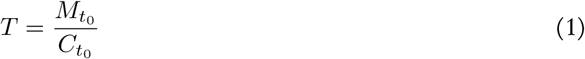

With 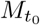: Number of deaths on that day *t*_0_;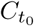: Number of cases reported on *t*_0_. We follow the reasoning of [4] that the fatality reported in each country can be used to create an index of the number of actual cases at a time *t*_0_. This index is :

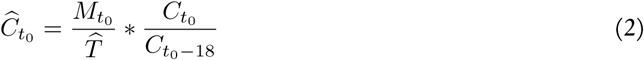

Where 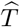: Estimated mortality rate; 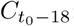: Number of cases reported in the country at time *t*_0_ minus 18 days. The proposed formula also makes it possible to evaluate the impact of policies to prevent the spread of the virus on the appearance of new cases. It is useful to more accurately estimate the number of people affected using a more consistent method for all countries [2]. We propose to follow a method developed in [4], to use the number of deaths reported by each country to estimate and compare the actual rate of affected people. This approach is based on four assumptions:

1. The mortality reported by each country is reliable
2. The fatality rate (*T*) is known and is similar in different countries
3. The average time between the onset of symptoms and death is known
4. The increase over time in the number of cases reported in the databases over the average time from symptom onset to death reflects the increase over time in the actual number of cases over the same period.

## 2 MATERIALS AND METHOD

We use a set of reported data to model the epidemic in Tunisia: data from the Tunisian Centre for Disease Control. It represents the epidemic transmission in Tunisia (see Figure 1). The first case was detected on March 2^*th*^ 2020. According to dataset *t* = 0 of the epidemic corresponds to 20 February.

**Figure 1.**
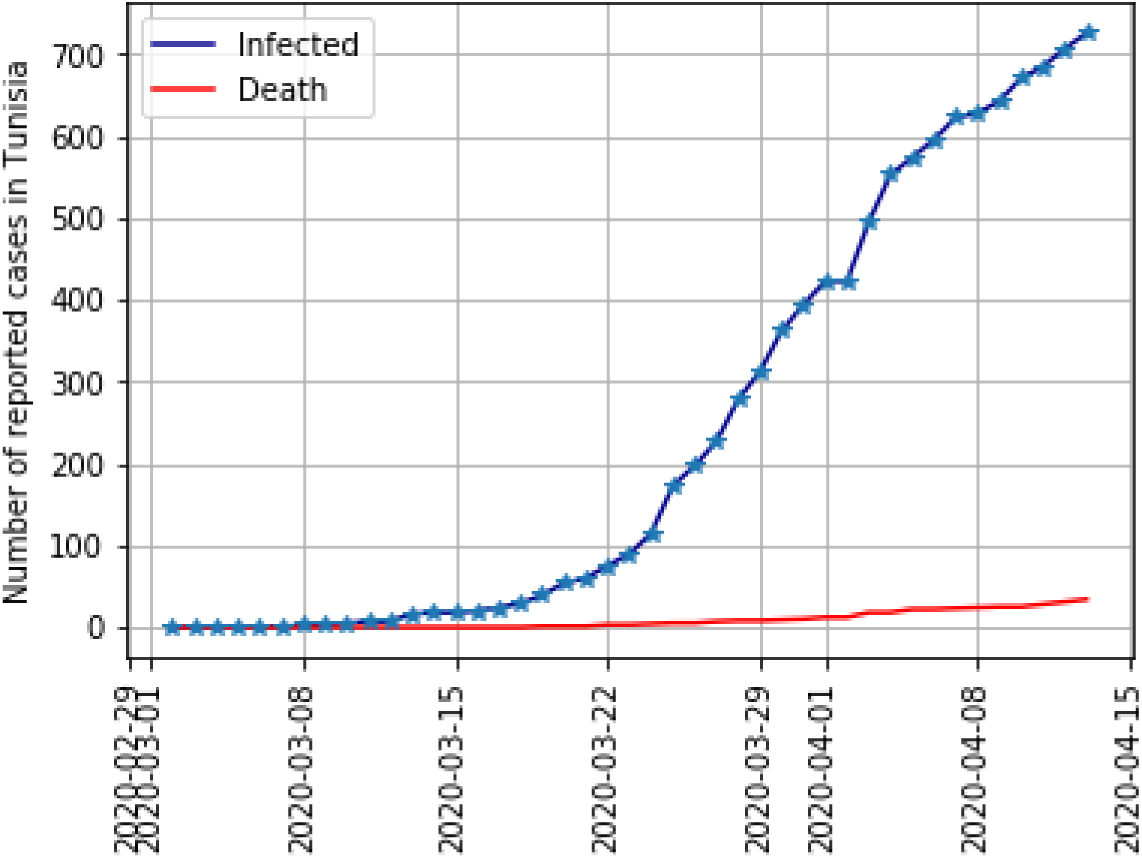
Infected cases and deaths in Tunisia

In [8] the epidemiological characteristics of patients are studied, and all patients who die on a given day have been infected much earlier, so the denominator of the fatality rate should be the total number of patients infected at the same time as those who died [9]. This is especially true as the rates of progression of the pandemic evolve differently in different countries: in April 2020, the number of people affected was increasing sharply from day to day in France, while it had stabilized in China. Figure 2 shows the reported mortality rate of COVID-19 in Tunisia.

**Figure 2.**
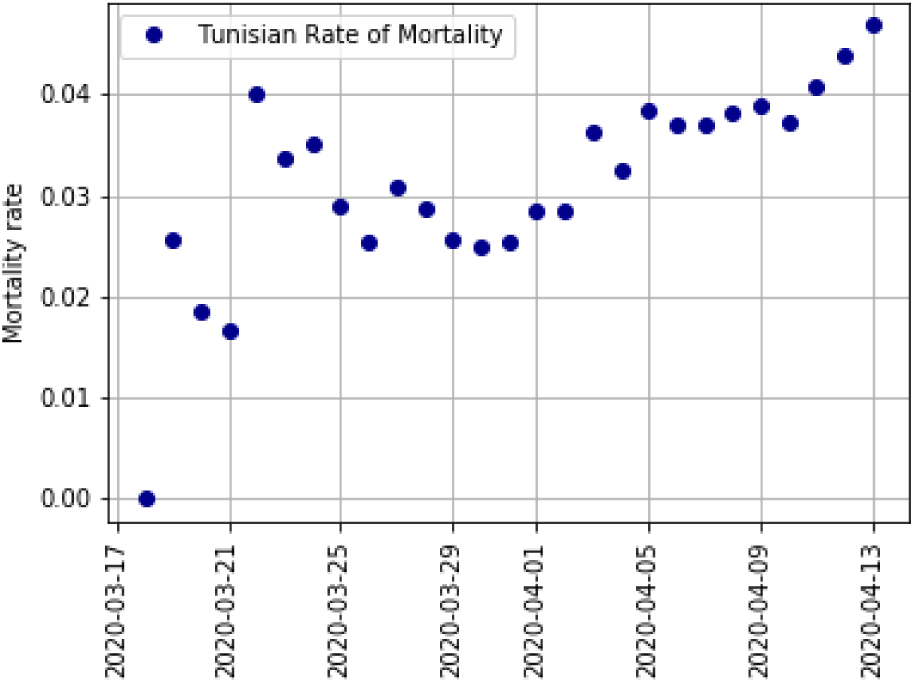
The mortality rate *T* in Tunisia varies from 1.67% to 4.68%. The first case of positive COVID-19 was declared in Tunisia on March 2^*th*^, 2020. The first case of death occurred on March 19^*th*^, 2020. The median of the fatality rate is equal to 3.26%

A better estimation of the fatality rate is therefore available:

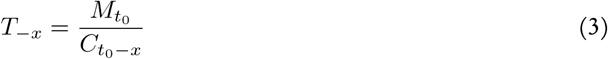

Where 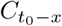 is the number of cases reported on day *t*_0_ minus *x* days, and *x* is the average time from symptom onset to death. A mean duration of 18 days is reported between the onset of symptoms of COVID-19 and patient death [10]. It has a 95% credible interval of 16.9 to 19.2 days [6]. An adjusted mortality rate *T*_*−*18_ can, therefore, be calculated by taking into account this average 18-day delay.

We can easily see in Figure 1 that the reported cases of the first two weeks were underestimated. Containment measures in Tunisia start on March 12 and become more restrictive on March 25. The median of *T*_*−*18_ in Figure 3 is equal to 0.67 and its mean is equal to 0.71. They are equal to 0.5 from March 30, 2020.

**Figure 3.**
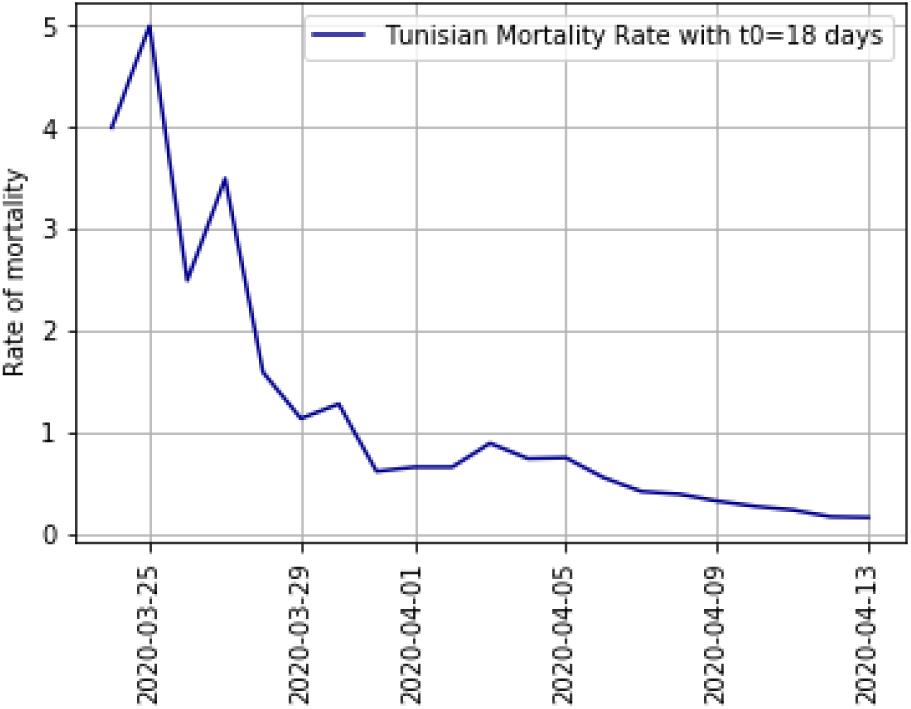
Tunisian mortality rate 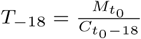 from March 24^*th*^ to April 13^*th*^, 2020

For *t*_0_ equal to 12 April 2020, Table 1 gives the main mortality rates. The computation takes into account the number of cases on March 26^*th*^, 2020. Data is available in [5].

**Table 1.**
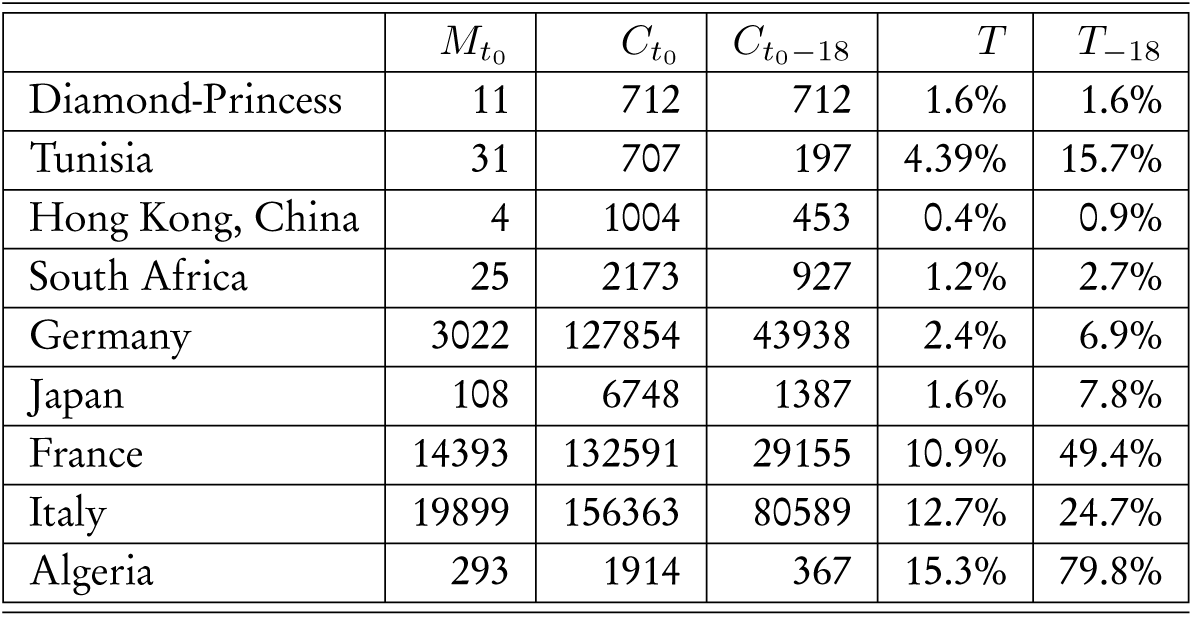
Fatality rates in different countries and in the cruise Diamond Princess

| | $M_{t_0}$ | $C_{t_0}$ | $C_{t_0-18}$ | $T$ | $T_{-18}$ |
| --- | --- | --- | --- | --- | --- |
| Diamond-Princess | 11 | 712 | 712 | 1.6% | 1.6% |
| Tunisia | 31 | 707 | 197 | 4.39% | 15.7% |
| Hong Kong, China | 4 | 1004 | 453 | 0.4% | 0.9% |
| South Africa | 25 | 2173 | 927 | 1.2% | 2.7% |
| Germany | 3022 | 127854 | 43938 | 2.4% | 6.9% |
| Japan | 108 | 6748 | 1387 | 1.6% | 7.8% |
| France | 14393 | 132591 | 29155 | 10.9% | 49.4% |
| Italy | 19899 | 156363 | 80589 | 12.7% | 24.7% |
| Algeria | 293 | 1914 | 367 | 15.3% | 79.8% |

## 3 RESULTS

With 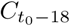 the number of cases reported on the day *t*_0_ minus 18 days. Values of *T*_*−*18_ in Table 1 reveal a great variation compared to *T* with variations ranging from 1% (Hong Kong, China) to 80% for Algeria.

The slope of this relationship varies greatly between countries, which is consistent with variable *T*_*−*18_. In [4] the values of *T*_*−*18_ based on cases reported by different countries are therefore unreliable. Several highly controlled international studies (mainland Chinese residents, returnees from mainland China, returnees from China, passengers on the Diamond-Princess cruise ship) reported *T*_*−*18_ values ranging from 0.7 to 3.6% [10]. Based on the latter study, an estimated mortality rate with 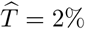 can be used. Knowing the number of deaths on day *t*_0_, the number of cases 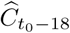 eighteen days before is estimated:

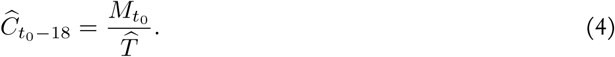

The progression of the number of cases reported over the last eighteen days *P*_18_ is known in each country. It depends on the rate of containment and its effectiveness. It is equal to:

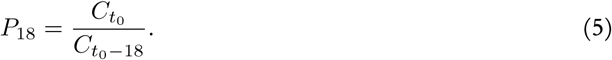

When comparing 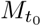 and 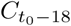 in different countries, [4] shows a linear relationship between mortality at *t*_0_ and the number of cases at *t*_0_ *−* 18 days for most of them. The number of cases estimated at *t*_0_ in each country is therefore

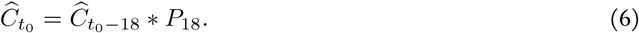

Values of 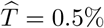 to 4% could be other suitable options. One of the limitations of our model is that this value may vary from country to country, for example depending on the distribution of the population into different age groups that have different sensitivity to COVID-19 [11]. Figure 4 shows the Tunisia cases’ estimation, where 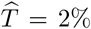 and 4%: The number of reported cases is underestimated. Note that some authors suggest that the real mortality rate for Covid-19 could be 5.6 to 15.6% [1], which is much higher values than those we used.

**Figure 4.**
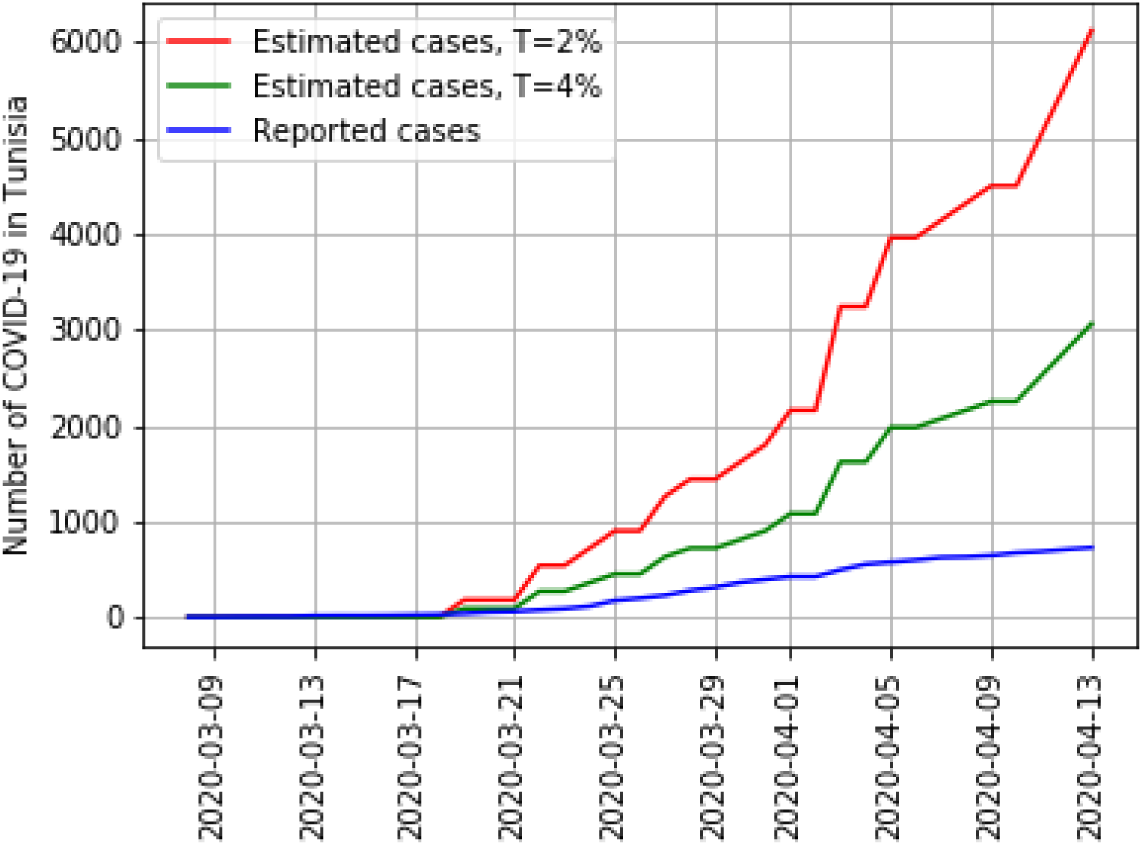
Estimation of 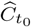, with death delay of 18 days, and 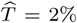 or 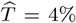. The number of estimated cases in Tunisia ranges from 2 790 and 5 580 at time *t*_0_ equals to April 12^*th*^, 2020

The population of Tunisia is equal to 11 818 619. For 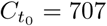 we obtain Table 3, which gives the evaluation of the number of cases in Tunisia as of April 12^*th*^, 2020 by modulating the estimated mortality rate 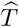 and taking into account delays of 12, 15 and 18 days between the onset of symptoms (or asymptomatic) and death [13]. We obtain *P*_12_ = 1.8, *P*_15_ = 2.5 and *P*_18_ = 3.6.

**Table 2.** Estimation of the number of cases in different countries as of April 12^*th*^, 2020 (*t*_0_) by correcting for the mortality rate taking into account a delay of 18 days between the onset of symptoms and death, and using 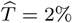

| | $M_{t_0}$ | $\hat{C}_{t_0-18}$ | $C_{t_0-18}$ | $C_{t_0}$ | $P_{18}$ | $\hat{C}_{t_0}$ |
| --- | --- | --- | --- | --- | --- | --- |
| Diamond-Princess | 11 | 550 | 712 | 712 | 1.0 | 550 |
| Tunisia | 31 | 1 550 | 197 | 707 | 3.6 | 5 580 |
| Hong Kong, China | 4 | 200 | 453 | 1 004 | 2.2 | 440 |
| South Africa | 25 | 1 250 | 927 | 2 173 | 2.3 | 2 875 |
| Germany | 3 022 | 151 100 | 43 938 | 127 854 | 2.9 | 438 190 |
| Japan | 108 | 5 400 | 1 387 | 6 748 | 4.9 | 26 460 |
| France | 14 393 | 719 650 | 29 155 | 132 591 | 4.5 | 3 238 425 |
| Italy | 19 899 | 994 950 | 80 589 | 156 363 | 1.9 | 1 890 405 |
| Algeria | 293 | 14 650 | 367 | 1 914 | 5.2 | 76 180 |

**Table 3.** Evaluation of the number of COVID-19 cases as of April 12^*th*^, 2020 by where the estimated mortality rate 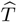 equals to 0.5%, 2%, 4%, 15%, and taking into account delays of 12, 15 and 18 days between the onset of symptoms (or asymptomatic) and death

|  | Simulation of cases in Tunisia at April 12<sup>th</sup>, 2020 |  |  |  |  |  |  |  |  |  |
| --- | --- | --- | --- | --- | --- | --- | --- | --- | --- | --- |
| Delay until death | 18 days |  |  |  | 15 days |  |  | 12 days |  |  |
| $\widehat{T}$ | 0.5% | 2% | 4% | 15% | 0.5% | 2% | 4% | 0.5% | 2% | 4% |
| $C_{t_0}$ | 22 320 | 5 580 | 2 790 | 744 | 15 500 | 3 875 | 1 937 | 11 160 | 2 790 | 1 395 |

It is conceivable that some people who died as a result of COVID-19 may not be counted. The number of infected people may be much higher than the number reported. Figure 5 estimates the number of cases in Tunisia as of April 12^*th*^, 2020 by modulating the estimated mortality rate 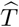 from 0.5 to 4% and taking into account delays of 12 days between the onset of symptoms (or asymptomatic) and death.

**Figure 5.**
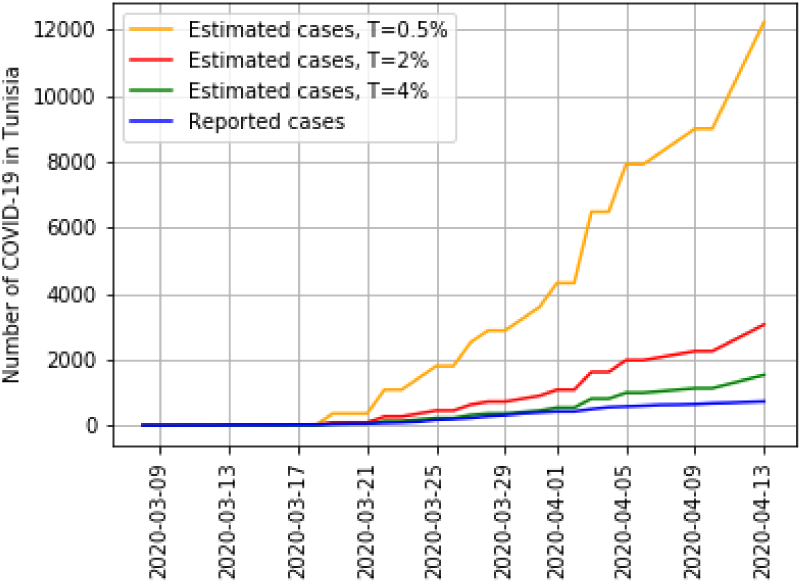
Estimated cases and reported cases of COVID-19 in Tunisia. Simulations using the average time, from first symptom to death, 12 days. The worse case is 11 160 and the optimistic case on April 12^*th*^, 2020 is 1 395

**Figure 6.**
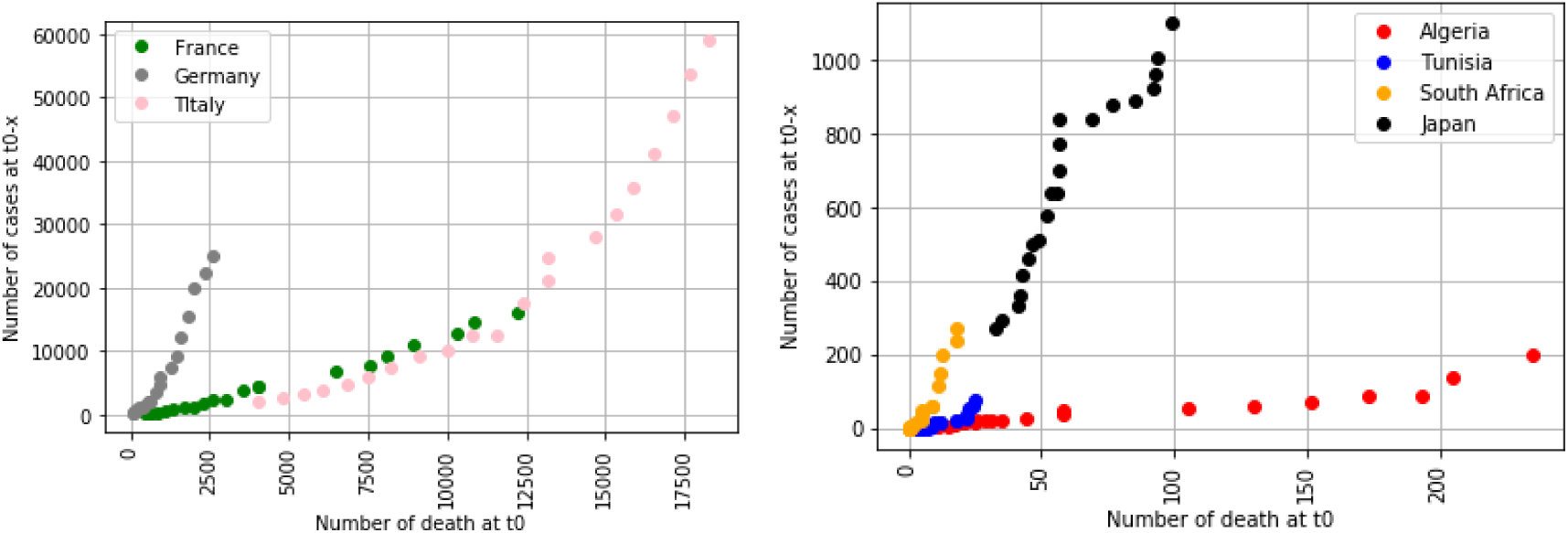
Number of deaths at *t*_0_ as a function of the number of cases eighteen days earlier (*t*_0_-18 days). These data are from March 2 to April 12^*th*^, 2020. The number of deaths in France, Italy, and Germany is greater than the number of deaths in Tunisia, Algeria, South Africa and Japan. These graphics show the relations between 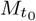 and 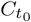

**Figure 7.**
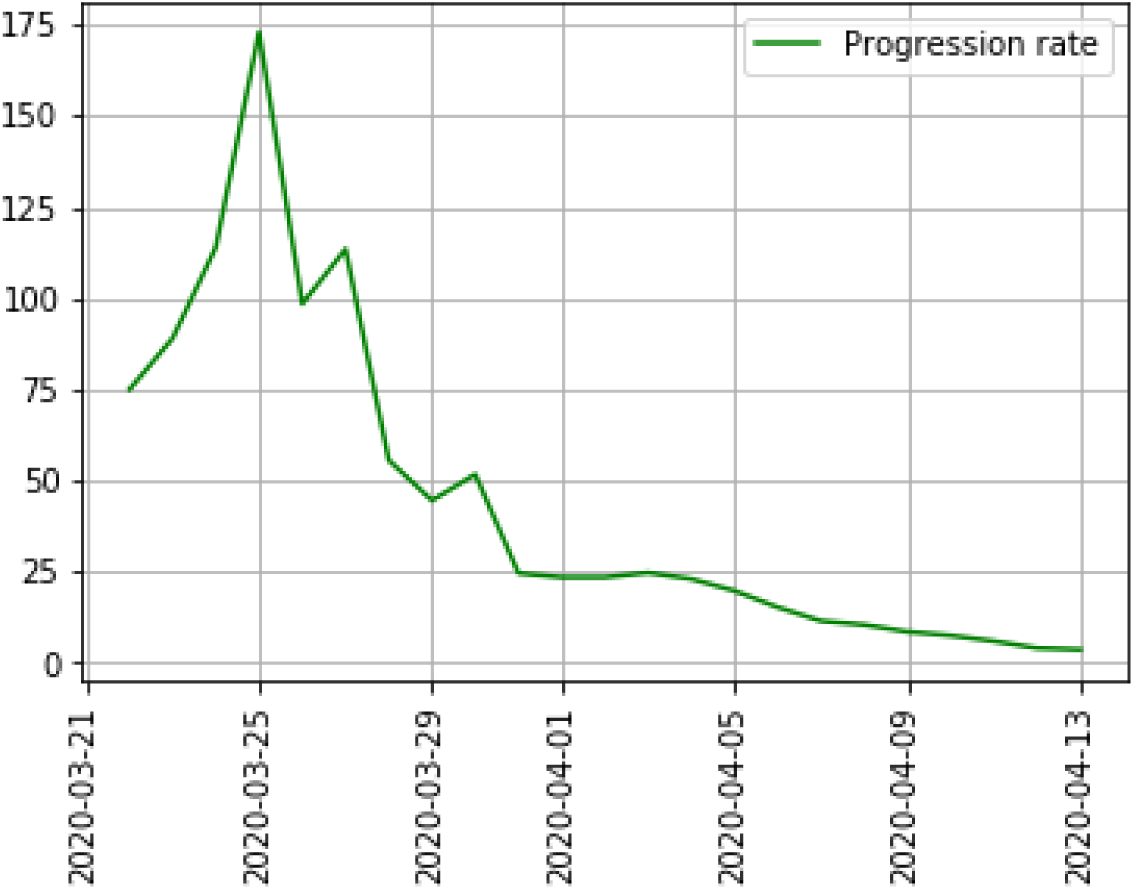
Effect of confinement measures on the progression rate *P*_18_ of COVID-19 in Tunisia. The progression rate *P*_18_ is equal to 173 on March 25 and decreases to 3.6 on April 12, 2020

**Figure 8.**
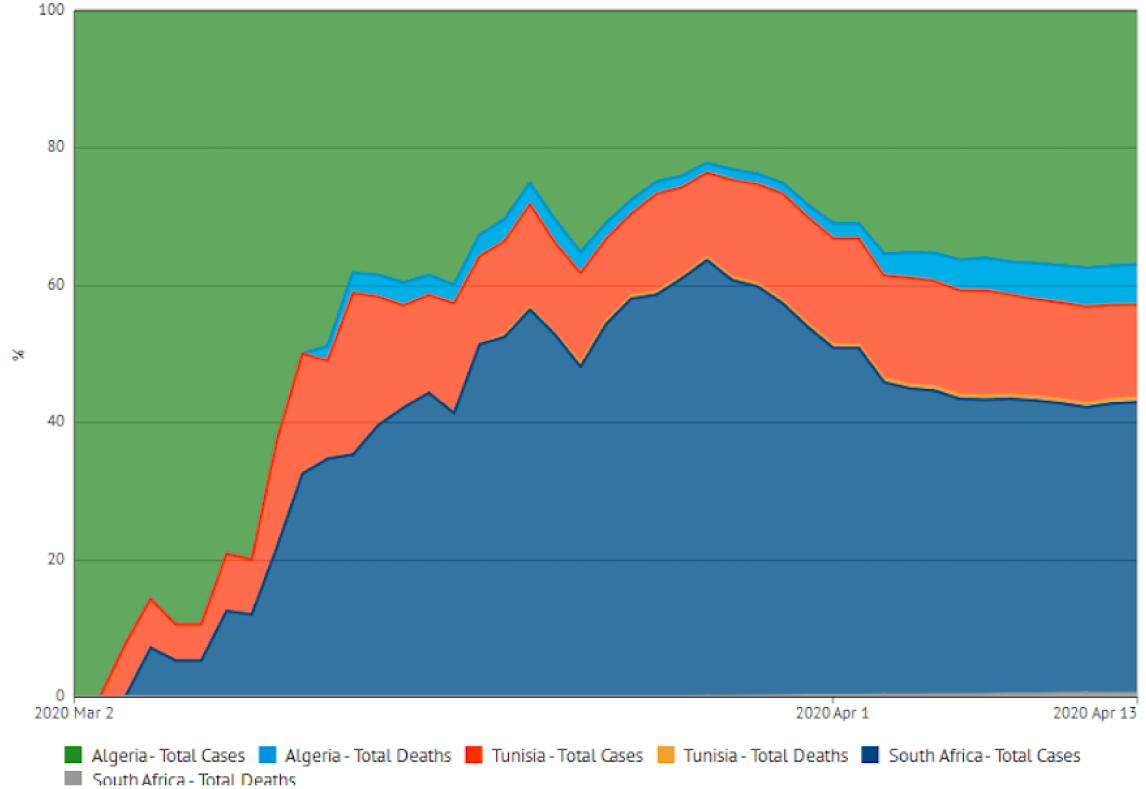
Reported cases of COVID-19 and deaths due to COVID-19 in Tunisia, Algeria and South Africa from March 2 to April 12^*th*^, 2020

## 4 DISCUSSION

This work uses an intuitive model, rather than other more complex models, such as [6] based on Bayesian approaches or [3]. As expected, our analysis suggests that the number of cases of COVID-19 in several countries exceeds the number of cases presented in international databases. Our results can contribute to the prevention and control of this epidemic in Tunisia. As a consequence of our study, we note that the number of cases is very sensitive. We can find multiple values of reported and unreported cases that provide a good fit for the data. Currently, there are many unanswered questions about this novel coronavirus, and this work enables us to clarify the effect of the mortality rate on the dynamic of the virus infection.

Public health measures, such as isolation, quarantine, and public closings, greatly reduce the final size of the epidemic and make the turning point much earlier than without these measures. For example, for Tunisia on April 12, *P*_18_ = 3.6, while on March 25, 2020, when regulation of progression rate based on population containment was still not fully efficient, *P*_18_ = 127. We can therefore estimate that the containment made it possible to reduce *P*_18_ from 127 to 3.6. The number of cases estimated on April 12 using *P*_18_ = 175 would have been 26 246 cases (with 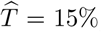). Thus, the containment from March 25 to April 12 prevented the appearance of 25 539 new cases in Tunisia, as well as associated fatalities.

An epidemic outbreak of a new human coronavirus COVID-19 will occur in Tunisia. For this outbreak, the unreported cases and the disease transmission rate are not identified. In order to recover these values from reported medical data, this simple mathematical model for estimating the COVID-19 cases is used. The knowledge of the cumulative reported symptomatic and asymptomatic infectious cases and assuming the infectious delay until death to be between 12 days and 18 days, this model estimates the number of infected cases. Then numerical simulations of the model are done to predict forward in time the severity of the epidemic, with adjusted data. We find that the most pessimistic number of infected people is 22 320 and the most optimistic is 744 at April 12^*th*^, 2020, and when the mortality rate is equal to 2%, the estimation number is equal to 5 580 (the number of reported cases at this date is 707 people).

This model inflated the number of cases, but it is at most a preliminary estimate. What is significant, in Tunisia and also in France, is that the public authorities are becoming aware of the necessity to properly quantify the number of deaths. This work raises interesting questions about the relation between the fatality rate and the number of infected cases. Our model used a mortality rate of 2% while several international studies reported rates of 0.7 to 3.6% [10]. Values from 0.5 to 4% could thus be other reasonable options to estimate mortality rate. One of the limitations of our model is that mortality rates can change from one country to another, depending on the distribution of the population in different age groups and on the co-morbidity that have different sensitivities to Covid-19. In the other hand, the ability of the virus to persist in different environments (hot climate) can affect this relation. Hence the importance of comparisons between countries with more and less sunshine, at different seasons and periods.

## Data Availability

All the data are public and available.

https://github.com/CSSEGISandData/COVID-19/tree/master/csse_covid_19_data/csse_covid_19_time_series

## ACKNOWLEDGMENTS

We would thank Khaled Abdeljaoued for his remarks following his careful reading of the paper.

## Appendix

The progression rate *P*_18_ shows the increase in the number of reported cases over the past eighteen days. Figure 7 proves the efficiency of confinement measure in Tunisia, where *P*_18_ decreases from March 25.

